# Neurodegeneration is strongly linked to heart failure severity and outcomes: framing the cardiocerebral syndrome

**DOI:** 10.1101/2023.09.15.23295547

**Authors:** Raphael Wurm, Suriya Prausmüller, Markus Ponleitner, Georg Spinka, Annika Weidenhammer, Henrike Arfsten, Gregor Heitzinger, Noel Gilian Panagiotides, Philipp Bartko, Georg Goliasch, Elisabeth Stögmann, Christian Hengstenberg, Martin Hülsmann, Noemi Pavo

## Abstract

**Background and Objectives:** Cognitive impairment is prevalent in patients with heart failure with reduced ejection fraction (HFrEF), affecting self-care and outcomes. Novel blood-based biomarkers have emerged as potential diagnostic tools for neurodegeneration. This study aimed to assess neurodegeneration in HFrEF by measuring neurofilament light chain (NfL), total tau (t-tau), amyloid-beta 42 (Aβ42), and 40 (Aβ40) in a large, well-characterised cohort.

**Methods:** The study included 470 HFrEF patients from a biobank-linked prospective registry at the Medical University of Vienna. High-sensitivity single-molecule assays were used for measurement. Unplanned hospitalisations and all-cause death were recorded as outcome parameters.

**Results:** All markers, but not the Aβ42/Aβ40 ratio, correlated with heart failure (HF) severity, i.e. NTproBNP and NYHA class, comorbidity burden and were significantly associated with all-cause death and HF-hospitalisations [crude HR for 1-log unit increase (95%CI): 4.44 (3.02-6.53), 5.04 (2.97-8-58), 3.90 (2.27-6.72) and 5.14 (2.84-9.32) for all-cause death and 2.48 (1.60-3.85), 3.44 (1.95-6.04), 3.13 (1.84-5.34) and 3.48 (1.93-6.27) for HHF, p<0.001 for all]. These markers remained significant after adjustment in multivariate models including NT-proBNP. NfL and t-tau showed the highest prognostic ability in the receiver operating characteristic analysis [AUC: 0.72, 0.68, 0.66, 0.67 for NfL, t-tau, Aβ40 and Aβ42, respectively]. The performance of NfL was comparable to that of NT-proBNP [C-index: 0.70 vs 0.72, p=0.225].

**Conclusions:** Neurodegeneration is directly interwoven with the progression of HF. Biomarkers, particularly NfL, may help identify patients profiting from detailed neurological workups. Further research is necessary to test if early diagnosis or optimised HFrEF treatment can preserve cognitive function.

## INTRODUCTION

Heart failure with reduced ejection fraction (HFrEF) can now be treated more effectively than ever, reducing cardiovascular events and extending life expectancy. Many patients with HFrEF now live well as septua- or octogenarians, where the preservation of cognitive function becomes increasingly crucial for enabling functional independence.

The prevalence of cognitive impairment (CI) and dementia steadily increases with age and rises sharply at the age of 80 years. The changes range from typical age-related cognitive changes to mild cognitive impairment and ultimately dementia, a condition characterised by cognitive dysfunction that hinders daily activities^1^. Risk factors for the development of CI are to some extent modifiable and include obesity, physical inactivity, diabetes, smoking, and hypertension^2^. A variety of factors may cause cumulative damage to the brain, but also neurodegenerative diseases result in the decline of cognitive function.

Degenerative dementias are the leading cause of significant cognitive decline in late life. Alzheimer’s disease (AD) is the most prevalent form of dementia, followed by vascular dementia and dementia with Lewy bodies^3^. Frequently multiple pathologies are present, but some affected individuals may not develop clinical dementia^4, 5^. AD is a condition characterised by extracellular aggregates of amyloid beta (Aβ) plaques and intracellular aggregations of neurofibrillary tangles, composed of hyperphosphorylated microtubule-associated tau^6^. These aggregates are neurotoxic, leading to neuronal loss and ultimately resulting in CI and dementia^7^.

Patients with HF frequently display decreased global cognitive function, exhibit deficits in multiple cognitive domains, and are at an increased risk of developing dementia^8^. This risk seems to increase with HF severity and duration of the disease. Furthermore, patients with HF and cognitive impairment tend to have poorer self-care and are at an increased risk for hospitalisations and death^9, 10^. HFrEF and CI indeed share some important risk factors^11^, yet the association was also reported to be independent, suggesting HF-specific mechanisms^8^. Impaired cerebral blood flow (CBF), a proinflammatory state, thromboembolic disease and protein abnormalities have been suggested to mediate this relationship^12^. Regarding amyloid pathophysiology and dementia, the reduction in cerebral CBF and the ensuing low-energy state in HFrEF may contribute to the formation of protein aggregates^13, 14^. Additionally, the hormonal response to a chronically low cardiac output entails endothelial dysfunction^15^. This together with reduced CBF disrupts the blood-brain-barrier^16^ and impedes clearance of Aβ and tau^17^. AD, on the other hand, might also increase the risk for development and worsening of HF. The cardiac tissue of patients with AD was found to contain Aβ similar to the deposits found in the brain, extending the concept from cerebral to systemic amyloidosis^18^.

CI is often mistakenly perceived as an appertaining bystander of HF and aging. To increase awareness and stimulate research, the condition was termed cardiocerebral syndrome (CCS)^12^. However, the current lack of definition, difficulties in making a definite diagnosis and understanding of the driving mechanisms hinder the development of effective diagnostic and therapeutic strategies. Recent developments in fluid biomarker assay technology employing Single-Molecule Arrays (Simoa) enable the detection of central nervous system (CNS)-based proteins secreted into the bloodstream with high sensitivity^19, 20^. Neurofilament light chain (NfL), total tau (t-tau), Aβ40 and Aβ42 can be determined on the Simoa platform.

This study aimed to explore the CCS by measuring blood-derived biomarkers of neurodegeneration and AD pathology, i.e. NfL, t-tau, Aβ40 and Aβ42, in a large population of unselected HFrEF patients in order to i) objectify neuronal damage, ii) investigate its relationship with HF severity and comorbidities, iii) show the association between neurodegenerative processes and HF outcomes, i.e. HF hospitalisations and mortality and to iv) compare the prognostic ability of the biomarkers of neurodegeneration.

## METHODS

### Study population

Chronic stable HfrEF patients were consecutively enrolled from a registry-biobank linkage at the Vienna General Hospital from February 2016 to January 2020. HFrEF diagnosis was established following the European Society of Cardiology guidelines, with patients undergoing guideline-directed therapy^21^. Data on comorbidities, clinical details, medication, and laboratory parameters were collected. Outcomes tracked included all-cause mortality, captured from the Austrian Death Registry, and unplanned HF-related hospitalizations from local and nationwide health records. The study followed the Declaration of Helsinki and received institutional ethics committee approval (EK1612/2015). All participants provided written consent.

### Blood sampling and routine laboratory analysis

Venous blood samples were collected during clinical visits for routine laboratory analysis, including N-terminal pro-brain natriuretic peptide (NT-proBNP), at the Medical University of Vienna’s Laboratory Medicine Department. Additional EDTA-plasma aliquots were stored at -80°C.

### Determination of plasma concentrations of NfL, t-tau, Aβ40 and Aβ40

EDTA plasma aliquots were used to measure NfL, t-tau, Aβ40, and Aβ42 concentrations. The Simoa NF-light kit (Quanterix, Lexington, MA, USA) was used to measure NfL, with a detection limit from 0 to ∼2000 pg/ml in plasma. The Simoa Human Neurology 3-Plex A assay (Quanterix) was used for t-tau, Aβ40, and Aβ42, with detection limits of 0 - ∼400 pg/ml, 0 - ∼600 pg/ml, and 0 - ∼200 pg/ml respectively. Procedures included sample thawing, calibration, dispensing, incubation, washing, and concentration analysis, all following manufacturer’s instructions. Sample readout and concentration analysis was performed with the Simoa SR-X Analyzer (Quanterix).

### Statistical analysis

Categorical and continuous variables were expressed as percentages and median with interquartile ranges (IQR), respectively, and compared using Fisher’s exact, Kruskal-Wallis and Mann-Whitney-U-tests. Correlations among biomarkers and clinical parameters were evaluated using Spearman’s rank correlation coefficient (r_s_). These biomarkers were also analyzed for their association with key HF comorbidities.

Cox proportional hazard regression was used to assess the association of log-transformed biomarkers with outcomes (hazard ratios and 95% CI). Multivariate analysis was conducted in three models, each progressively adding confounders (see Table 3). The association of these biomarkers with outcomes was also illustrated using restricted cubic spline curves, and predictive performance was evaluated by ROC curves and Harrell’s C-indices.

To investigate the association of neurodegeneration markers with critical clinical parameters in HFrEF, a linear regression model was constructed. Predictors, selected based on a stepwise forward approach, included demographics, comorbidities, and disease severity-related lab parameters (see Supplementary Table 1).

All analyses were two-sided with a significance level of 0.05, performed using SPSS version 24 and R version 4.2.2.

## RESULTS

### Study population

A total of 470 HFrEF patients were included in the study. Table 1 displays the detailed baseline characteristics of the study population. The median age was 62 years (Q1-Q3: 52-72) and 363 (77%) patients were male. NT-proBNP concentration was elevated with a median of 1812 pg/ml (IQR: 718–3881). Most patients were in NYHA functional class II (47%) or class III (39%). Guideline-directed medical therapy was well established as 439 (93%) patients received renin-angiotensin-aldosterone-system inhibitors (RASi), 444 (95%) patients were treated with beta-blockers and 355 (76%) patients with mineralocorticoid receptor-antagonists. Baseline plasma levels of Nfl, t-tau, Aβ40, Aβ42 and Aβ42/Aβ40 ratio were 25.6 pg/ml (Q1-Q3: 14.7-45.8), 1.01 (Q1-Q3: 0.63-1.56), 183.9 pg/ml (Q1-Q3: 106.2-272.5), 8.53 pg/ml (Q1-Q3: 5.37-11.82) and 0.048 pg/ml (Q1-Q3: 0.039-0.057), respectively.

**Table 1.**
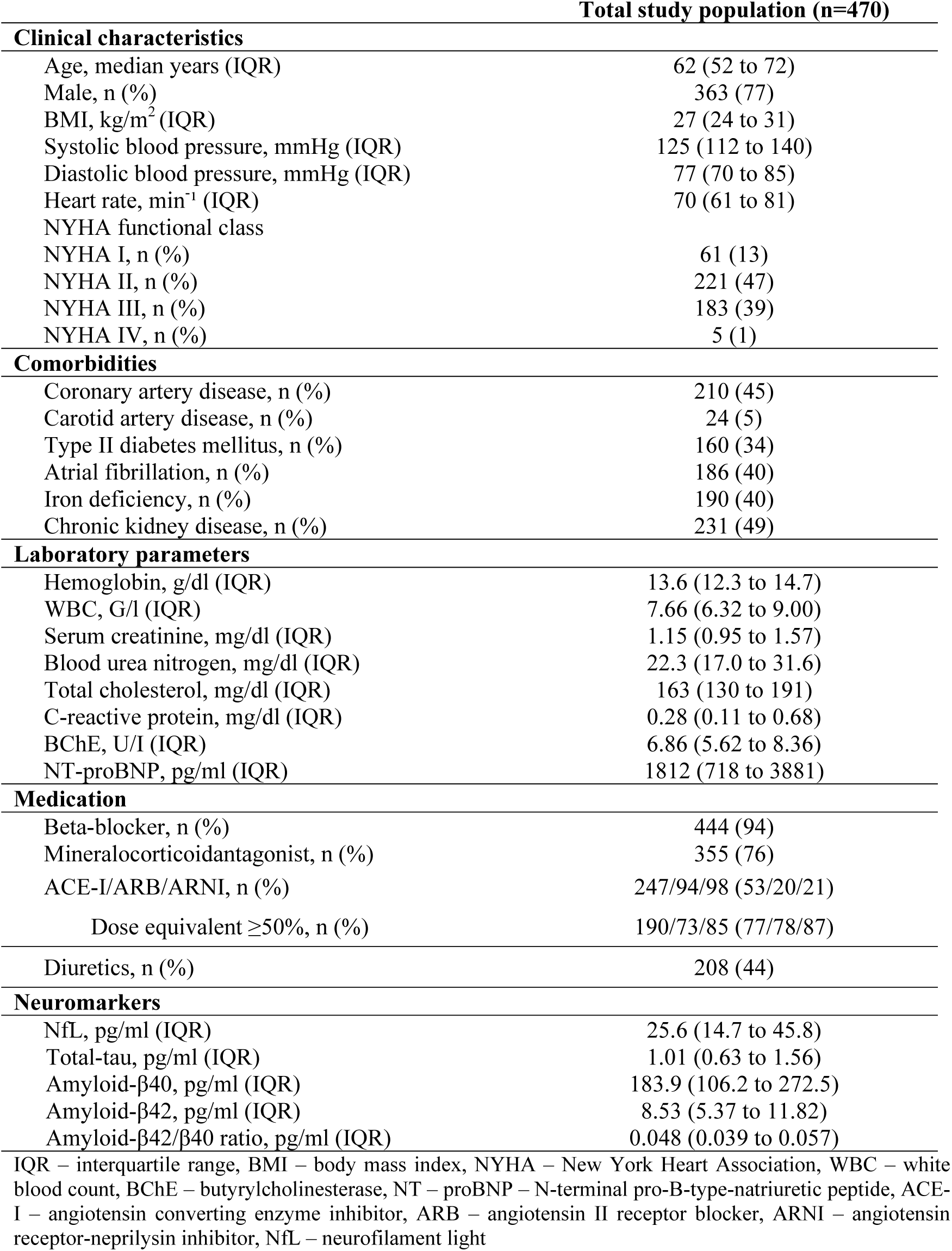
Baseline characteristics of the HFrEF study population. Continuous variables are given as medians and interquartile ranges (IQR), counts are given as numbers and percentages.

| Total study population (n=470) |  |
| --- | --- |
| **Clinical characteristics** |  |
| Age, median years (IQR) | 62 (52 to 72) |
| Male, n (%) | 363 (77) |
| BMI, kg/m<sup>2</sup> (IQR) | 27 (24 to 31) |
| Systolic blood pressure, mmHg (IQR) | 125 (112 to 140) |
| Diastolic blood pressure, mmHg (IQR) | 77 (70 to 85) |
| Heart rate, min<sup>-1</sup> (IQR) | 70 (61 to 81) |
| NYHA functional class |  |
| NYHA I, n (%) | 61 (13) |
| NYHA II, n (%) | 221 (47) |
| NYHA III, n (%) | 183 (39) |
| NYHA IV, n (%) | 5 (1) |
| **Comorbidities** |  |
| Coronary artery disease, n (%) | 210 (45) |
| Carotid artery disease, n (%) | 24 (5) |
| Type II diabetes mellitus, n (%) | 160 (34) |
| Atrial fibrillation, n (%) | 186 (40) |
| Iron deficiency, n (%) | 190 (40) |
| Chronic kidney disease, n (%) | 231 (49) |
| **Laboratory parameters** |  |
| Hemoglobin, g/dl (IQR) | 13.6 (12.3 to 14.7) |
| WBC, G/l (IQR) | 7.66 (6.32 to 9.00) |
| Serum creatinine, mg/dl (IQR) | 1.15 (0.95 to 1.57) |
| Blood urea nitrogen, mg/dl (IQR) | 22.3 (17.0 to 31.6) |
| Total cholesterol, mg/dl (IQR) | 163 (130 to 191) |
| C-reactive protein, mg/dl (IQR) | 0.28 (0.11 to 0.68) |
| BChE, U/l (IQR) | 6.86 (5.62 to 8.36) |
| NT-proBNP, pg/ml (IQR) | 1812 (718 to 3881) |
| **Medication** |  |
| Beta-blocker, n (%) | 444 (94) |
| Mineralocorticoid antagonist, n (%) | 355 (76) |
| ACE-I/ARB/ARNI, n (%) | 247/94/98 (53/20/21) |
| Dose equivalent ≥50%, n (%) | 190/73/85 (77/78/87) |
| Diuretics, n (%) | 208 (44) |
| **Neuromarkers** |  |
| NfL, pg/ml (IQR) | 25.6 (14.7 to 45.8) |
| Total-tau, pg/ml (IQR) | 1.01 (0.63 to 1.56) |
| Amyloid-β40, pg/ml (IQR) | 183.9 (106.2 to 272.5) |
| Amyloid-β42, pg/ml (IQR) | 8.53 (5.37 to 11.82) |
| Amyloid-β42/β40 ratio, pg/ml (IQR) | 0.048 (0.039 to 0.057) |

IQR – interquartile range, BMI – body mass index, NYHA – New York Heart Association, WBC – white blood count, BChE – butyrylcholinesterase, NT – proBNP – N-terminal pro-B-type-natriuretic peptide, ACE-I – angiotensin converting enzyme inhibitor, ARB – angiotensin II receptor blocker, ARNI – angiotensin receptor-neprilysin inhibitor, NfL – neurofilament light

### Association of NfL, t-tau and amyloid β with clinical and laboratory parameters

The correlation table between the neurodegenerative biomarkers and clinical and laboratory parameters are provided as Supplementary Figure 1. All neurodegenerative markers showed a correlation with age, the strongest for NfL (r_s_=0.45, p<0.001) and only a weak correlation for t-tau, Aβ40 and Aβ42, and no correlation with the Aβ42/Aβ40 ratio. No difference was observed for neuromarker distributions according to sex. Solely NfL displayed an inverse correlation with BMI (r_s_=-0.18, p<0.001). NfL, t-tau, Aβ40 and Aβ42 levels showed a direct correlation with creatinine and urea (creatinine: r_s_= 0.53; r_s_=0.37; r_s_=0.39; r_s_=0.39; urea: r_s_= 0.49; r_s_=0.32; r_s_=0.31; r_s_=0.33; respectively, p<0.001 for all). Neuromarkers correlated inversely with hemoglobin, and butyrylcholinesterase but less with aspartate aminotransferase and alanine transaminase, and weakly with C-reactive protein. Neither lipids nor electrolytes showed a remarkably strong correlation with the neuromarkers.

### Association of NfL, tau and amyloid β with HF severity and comorbidities

The relationship between NfL, t-tau, Aβ40, Aβ42 and the Aβ42/40 ratio and HF severity is depicted in Figure 1A. NfL, tau, Aβ40 and Aβ42 concentrations showed a significant direct correlation with NT-proBNP (r_s_=0.436, p<0.001; r_s_=0.389, p<0.001; r_s_=0.420, p<0.001 and r_s_=0.376, p<0.001, respectively), whereas the Aβ42/Aβ40 ratio showed only a weak inverse correlation (r_s_=-0.154, p<0.001). NfL, t-tau, Aβ40 and Aβ42, but not the Aβ42/Aβ40 ratio, were also significantly and directly associated with NYHA class (p_trend_<0.001 for all). Figure 1B illustrates the association between the neuromarkers and main comorbidities. Supplementary Figure 2 depicts levels of the neuromarkers according to the comorbidity burden. All neuromarkers were elevated in individuals with CKD and known carotid artery stenosis (NfL: p<0.001 for both; t-tau: p<0.001 and p=0.041; Aβ40: p<0.001 and p=0.050; Aβ42: p<0.001 and p=0.033, respectively). In patients with T2DM and CAD, only NfL levels were significantly elevated (p<0.001 for both), whereas atrial fibrillation was associated with higher levels of NfL and t-tau (p<0.001 for both). In individuals with ID, only Aβ42 levels were significantly higher (p=0.025). No significant association was observed between comorbidities and the Aβ42/Aβ40 ratio.

**Figure 1.**
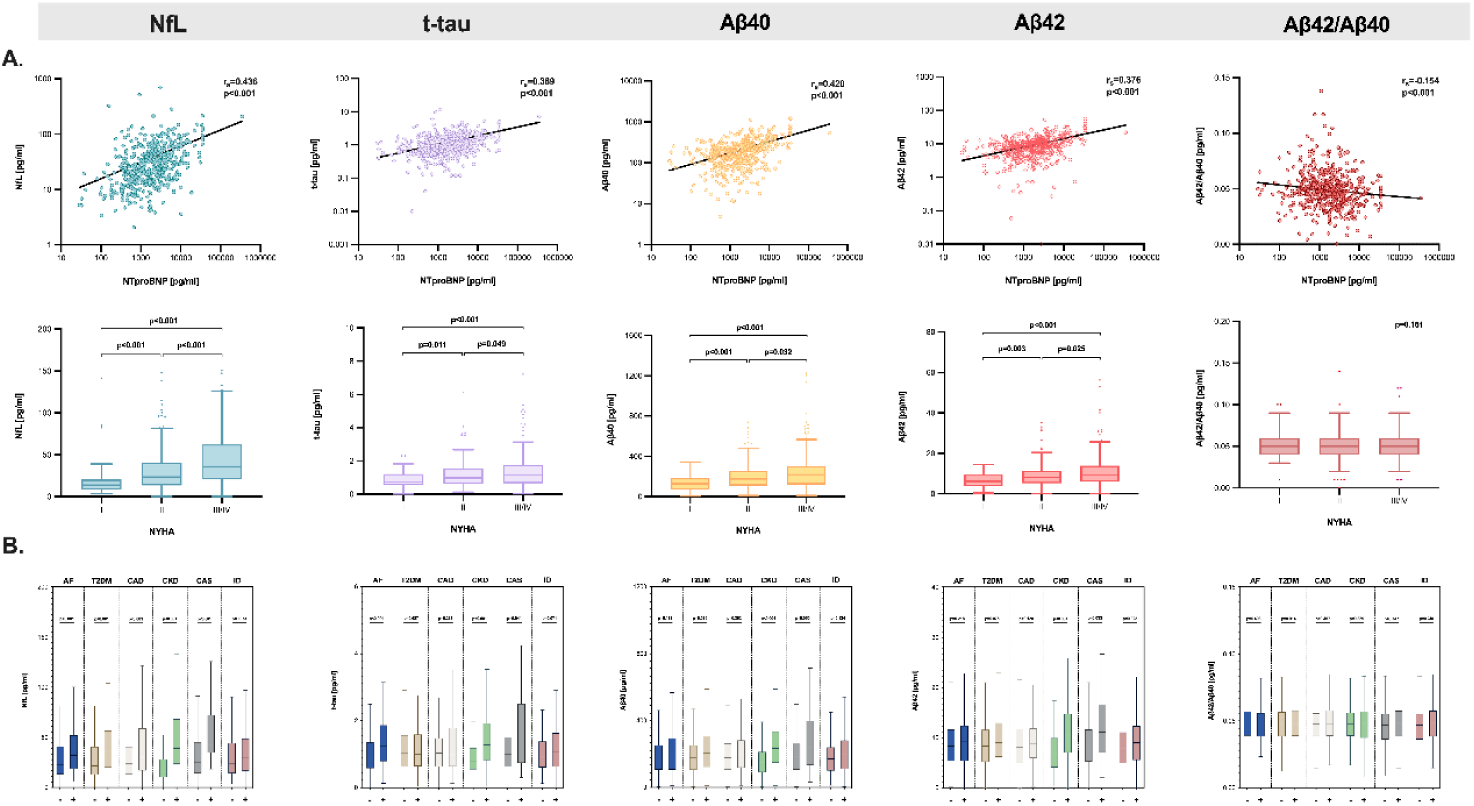
Relationship of the neuromarkers neurofilament light chain (NfL), total tau protein (t-tau), amyloid β40, amyloid β42 and the amyloid β42/40 ratio with heart failure severity and comorbidities. A. Scatter plots with linear regression analysis and the Spearman rho coefficient are shown for the association between neuromarkers and NT-proBNP. Tukey-boxplots and group comparisons are shown for the association between neuromarkers and (A.) New York Heart Association (NYHA) classes and (B.) the comorbidities atrial fibrillation (AF), type 2 diabetes mellitus (T2DM), coronary artery disease (CAD), chronic kidney disease (CKD), carotid artery stenosis (CAS) and iron deficiency (ID). Comparisons were made by the Mann-Whitney test and p-values are indicated in the graph. B. Cubic spline analysis for the respective parameters regarding all-cause mortality. The distribution of NfL, tau, amyloid β40, amyloid β42 and the amyloid β42/40 ratio are indicated below the curves.

### Influence of patients’ characteristics on neuromarker levels

The results of the multivariate regression analysis are presented in Table 2. Overall, the goodness of fit, as indicated by the R^2^ values, was modest for all markers of neurodegeneration (R^2^: NfL: 0.17, t-tau: 0.20, Aβ40: 0.36, Aβ42: 0.36 and Aβ42/Aβ40 ratio: 0.05). NT-proBNP and eGFR emerged as significant predictors for all neurodegeneration markers, except for the Aβ42/Aβ40 ratio. Aβ40 and Aβ42 levels were additionally associated with age and C-reactive protein. Among the comorbidities, only a history of hypertension was found to be related to t-tau levels.

**Table 2.**
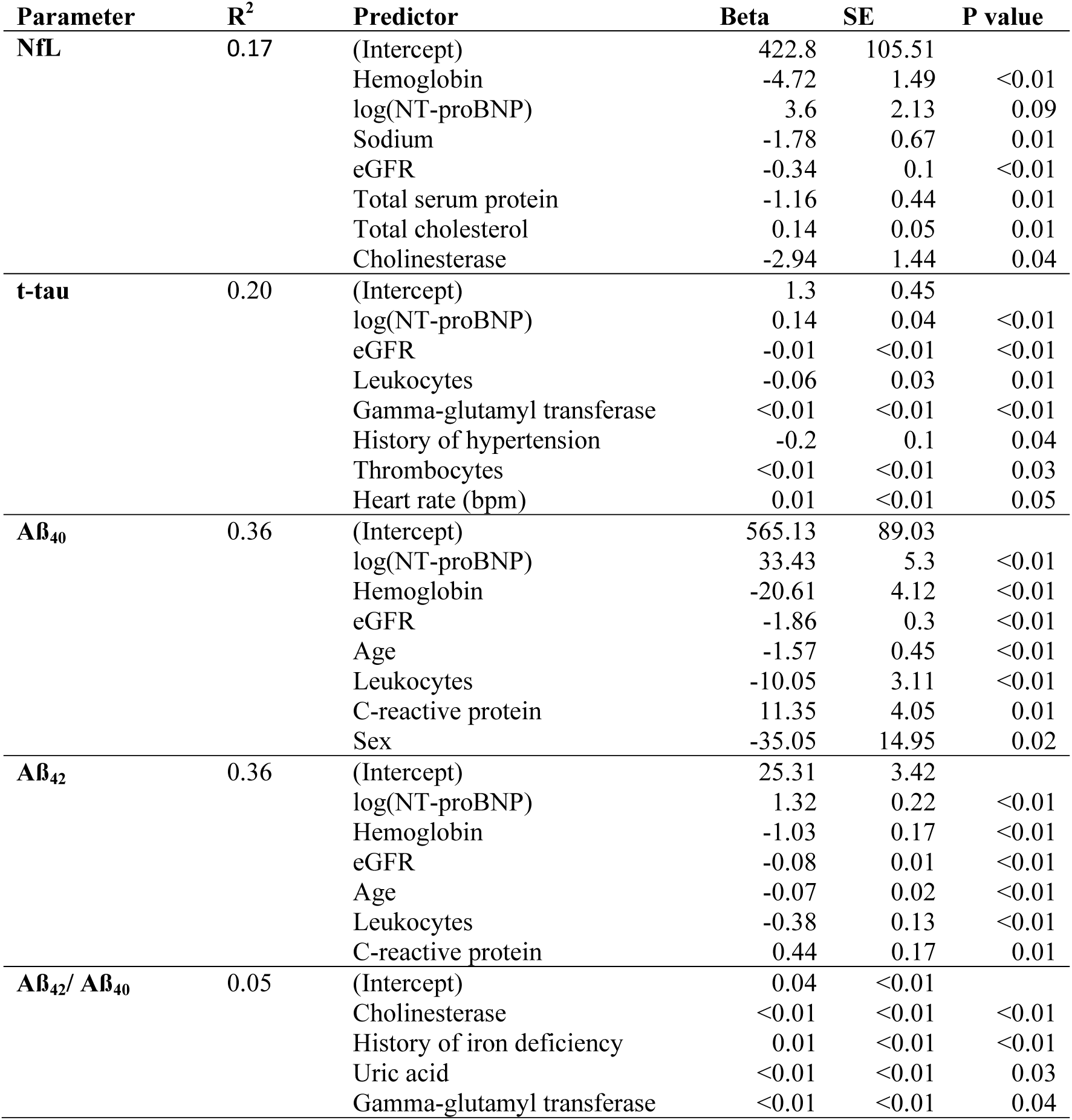
Linear regression model with NfL, t-tau, Aß40 and Aß42 as the dependent variables. Parameters to be entered in the model were selected with a stepwise forward approach based on p-values (inclusion: <0.05, exclusion: >0.10). The first parameter to enter the model is the independent variable with the lowest p-value.

| Parameter | R<sup>2</sup> | Predictor | Beta | SE | P value |
| --- | --- | --- | --- | --- | --- |
| **NfL** | 0.17 | (Intercept) | 422.8 | 105.51 |  |
|  |  | Hemoglobin | -4.72 | 1.49 | <0.01 |
|  |  | log(NT-proBNP) | 3.6 | 2.13 | 0.09 |
|  |  | Sodium | -1.78 | 0.67 | 0.01 |
|  |  | eGFR | -0.34 | 0.1 | <0.01 |
|  |  | Total serum protein | -1.16 | 0.44 | 0.01 |
|  |  | Total cholesterol | 0.14 | 0.05 | 0.01 |
|  |  | Cholinesterase | -2.94 | 1.44 | 0.04 |
| **t-tau** | 0.20 | (Intercept) | 1.3 | 0.45 |  |
|  |  | log(NT-proBNP) | 0.14 | 0.04 | <0.01 |
|  |  | eGFR | -0.01 | <0.01 | <0.01 |
|  |  | Leukocytes | -0.06 | 0.03 | 0.01 |
|  |  | Gamma-glutamyl transferase | <0.01 | <0.01 | <0.01 |
|  |  | History of hypertension | -0.2 | 0.1 | 0.04 |
|  |  | Thrombocytes | <0.01 | <0.01 | 0.03 |
|  |  | Heart rate (bpm) | 0.01 | <0.01 | 0.05 |
| **Aβ<sub>40</sub>** | 0.36 | (Intercept) | 565.13 | 89.03 |  |
|  |  | log(NT-proBNP) | 33.43 | 5.3 | <0.01 |
|  |  | Hemoglobin | -20.61 | 4.12 | <0.01 |
|  |  | eGFR | -1.86 | 0.3 | <0.01 |
|  |  | Age | -1.57 | 0.45 | <0.01 |
|  |  | Leukocytes | -10.05 | 3.11 | <0.01 |
|  |  | C-reactive protein | 11.35 | 4.05 | 0.01 |
|  |  | Sex | -35.05 | 14.95 | 0.02 |
| **Aβ<sub>42</sub>** | 0.36 | (Intercept) | 25.31 | 3.42 |  |
|  |  | log(NT-proBNP) | 1.32 | 0.22 | <0.01 |
|  |  | Hemoglobin | -1.03 | 0.17 | <0.01 |
|  |  | eGFR | -0.08 | 0.01 | <0.01 |
|  |  | Age | -0.07 | 0.02 | <0.01 |
|  |  | Leukocytes | -0.38 | 0.13 | <0.01 |
|  |  | C-reactive protein | 0.44 | 0.17 | 0.01 |
| **Aβ<sub>42</sub>/ Aβ<sub>40</sub>** | 0.05 | (Intercept) | 0.04 | <0.01 |  |
|  |  | Cholinesterase | <0.01 | <0.01 | <0.01 |
|  |  | History of iron deficiency | 0.01 | <0.01 | <0.01 |
|  |  | Uric acid | <0.01 | <0.01 | 0.03 |
|  |  | Gamma-glutamyl transferase | <0.01 | <0.01 | 0.04 |

**Table 3.**
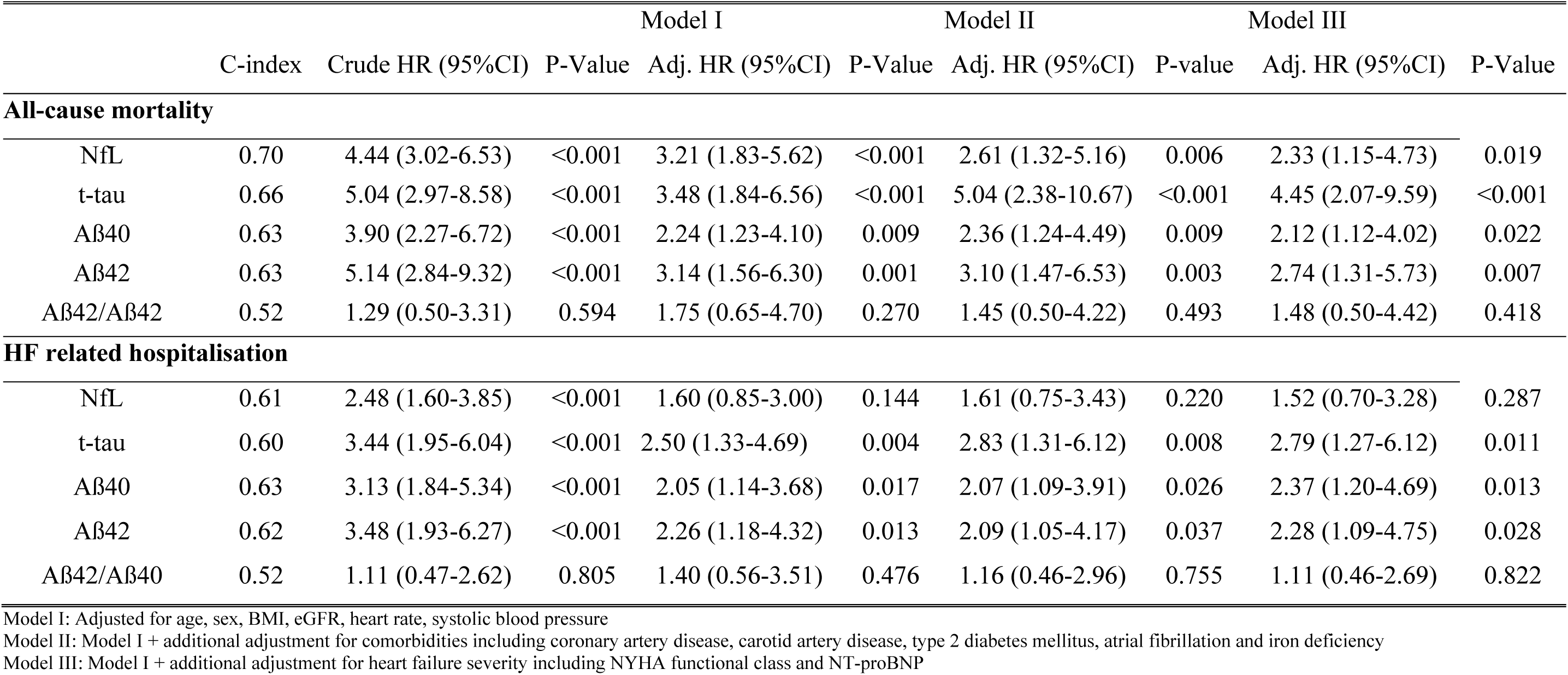
Association of biomarkers of neurodegeneration with HF outcomes. C-Index, crude and multivariate Cox regression analysis for romarkers neurofilament light chain (NfL), total tau protein (t-tau), amyloid β40, amyloid β42 and the amyloid β42/40 ratio with regards to all-se mortality and HF-related hospitalisations are shown. Hazard ratio (HR) is expressed for 1-log unit increase.

### Association of of NfL, tau and amyloid β with outcome

After a median follow-up of 3.7 years (Q1-Q3: 2.5-4.9), 144 (31%) patients died. Unplanned HF-related rehospitalisations occurred in 135 (29%) patients with a median follow-up time of 1.08 years (Q1-Q3: 0.3-2.9). Table 3 presents the results of the Cox regression analysis. Increased levels of NfL, t-tau, Aβ40 and Aβ42 were predictive for worse overall survival in the univariate model (crude HR for an increase in 1-log unit: 4.44 [95%CI: 3.02 to 6.53], 5.04 [95%CI: 2.97 to 8.58], 3.90 [95%CI: 2.27 to 6.72], and 5.14 [95%CI: 2.84 to 9.32], respectively; p<0.001 for all). The results remained virtually unchanged after adjustment for potential confounders. In contrast, the Aβ42/40 ratio showed no significant association with all-cause mortality (crude HR Aβ42/40 ratio for an increase in 1-log unit: 1.29 [95%CI: 0.50 to 3.31], p=0.594).

In terms of HF-related hospitalisations, elevated levels of NfL, t-tau, Aβ40, and Aβ42 were similarly all significantly associated with an increased risk (crude HR for an increase in 1-log unit: 2.48 [95%CI: 1.60 to 3.85], 3.44 [95%CI: 1.95 to 6.04], 3.13 [95%CI: 1.84 to 5.34], and 3.48 [95%CI: 1.93 to 6.27], respectively; p<0.001 for all) in the univariate analysis. After adjusting for various confounders t-tau, Aβ40, and Aβ42 remained predictors for HF-related hospitalisations. The Aβ42/Aβ40 ratio did not show a significant association with HF-related hospitalisations (crude HR for an increase in 1-log unit: 1.11, 95% CI: 0.47-2.62, p=0.805). Cubic spline modeling showed a steady increase in mortality and HF hospitalisation risk with an increase in serum. levels for all neuromarkers, except the ratio, with cut-offs of approximately NfL 20pg/ml, t-tau 1.4pg/ml, Aβ40 200pg/ml and Aβ42 10pg/ml (Figure 2).

**Figure 2.**
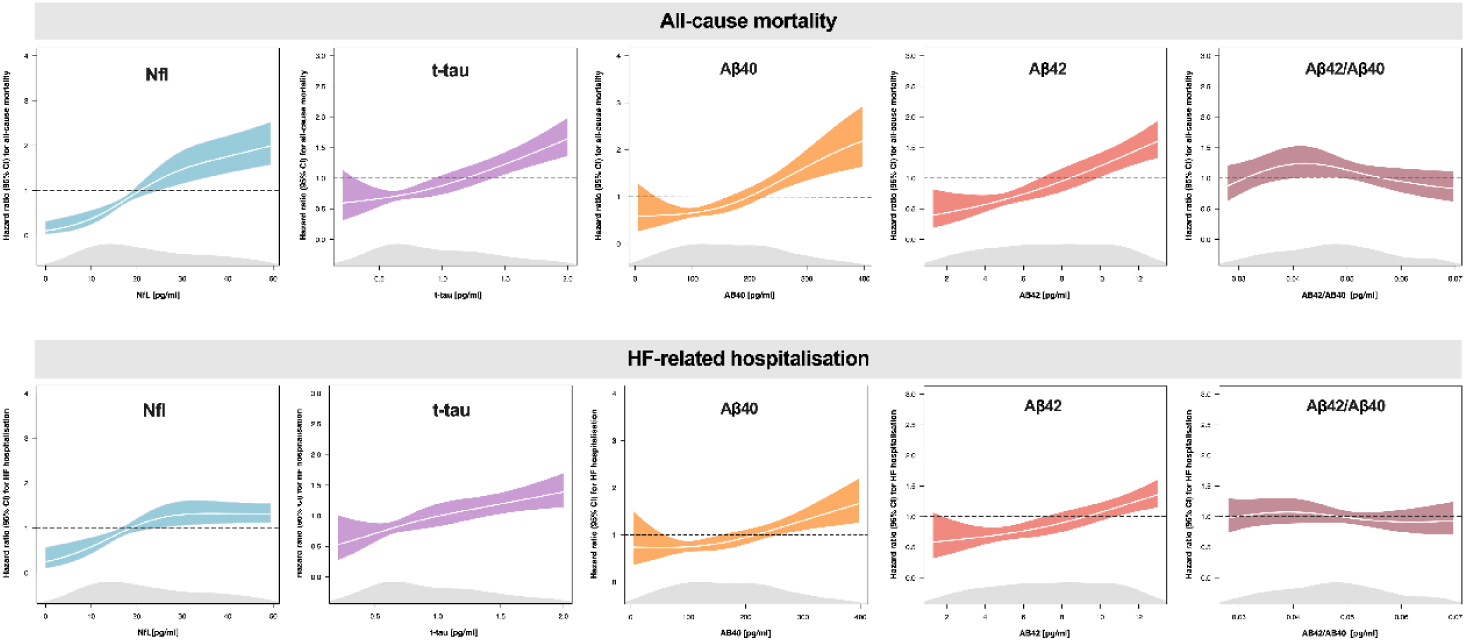
Cubic spline analysis for the respective parameters regarding all-cause mortality and unplanned HF-related hospitalisations. The distribution of NfL, t-tau, amyloid β40, amyloid β42 and the amyloid β42/40 ratio are indicated below the curves.

The results of the C-statistics and ROC analysis are presented in Table 3 and Figure 3. The discriminatory accuracy of NfL in predicting all-cause mortality was comparable to the well-established risk marker NT-proBNP (C-index: 0.70 vs. 0.72, p=0.225), while the C-indices of t-tau, Aβ40, Aβ42 and the Aβ42/40 ratio were significantly lower compared to NT-proBNP (C-index: 0.66 vs. 0.72, p=0.034, 0.63 vs. 0.72, p=0.003, 0.63 vs. 0.72, p=0.003, 0.52 vs. 0.72, p<0.001; respectively). In terms of predicting HF-related hospitalisations, NfL, t-tau, Aβ40 and Aβ42 performed similarly to NT-proBNP (C-index: 0.61 vs. 0.68, p=0.103; 0.60 vs. 0.68, p=0.089; 0.63 vs. 0.68, p=0.153; 0.62 vs. 0.68, p=0.140; respectively). Aβ42/40 ratio showed insufficient performance for predicting all-cause mortality and all HF-hospitalisations (C-index: 0.52 for both endpoints).

**Figure 3.**
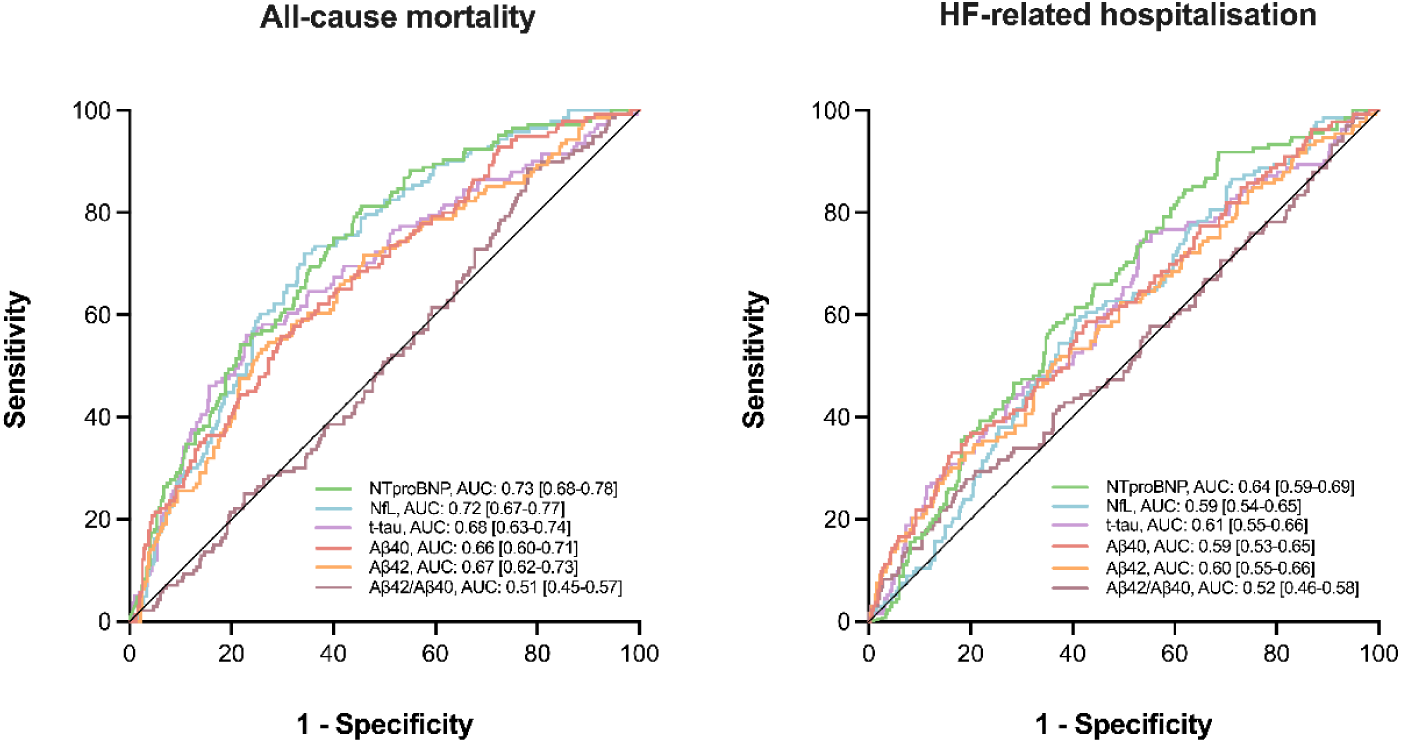
Receiver operating characteristic (ROC) curves for the endpoint all-cause mortality and unplanned heart failure related hospitalisations for the neuromarkers neurofilament light chain (NfL), total tau protein (t-tau), Aβ40, Aβ42, Aβ42/ Aβ40 ratio and the risk marker NT-proBNP.

## DISCUSSION

The study presents results from a comprehensive investigation of specific biomarkers of neurodegeneration and Aβ pathology in a cohort of HFrEF patients on guideline-directed medical therapy. The data show that a neurodegenerative process, evidenced by levels of NfL and t-tau, is readily detectable and that biomarker levels are elevated in HF compared to healthy individuals reported in previous studies. NfL, t-tau, Aβ40 and Aβ42 concentrations, but not the ratio of Aβ42/40, are significantly associated with the severity of disease, mirrored by NT-proBNP and NYHA class, and comorbidities, especially T2DM. The increase in NfL, t-tau, Aβ40 and Aβ42 is associated with worse overall survival and an increased risk of HF hospitalisations, independently from NT-proBNP or renal function. Notably, NfL displayed a comparable prognostic ability to NT-proBNP.

### Markers of neurodegeneration NfL, t-tau and amyloid pathology

Blood biomarkers hold promise for future diagnostic approaches in neurodegenerative disease as they are more scalable than biomarkers of the cerebrospinal fluid and brain imaging markers. They similarly offer the exploration of a possible neuronal injury process associated with HF.

#### NfL

NfL is a cytoplasmic protein expressed in neurons of the central and peripheral nervous system. It is involved in maintaining the structural integrity of dendrites, axons, and soma. It is released into the bloodstream in many conditions with neuronal damage. Emerging data indicate the clinical utility of blood-based measurements of NfL in neurodegenerative diseases. NfL has recently been shown to be of diagnostic value for AD or multiple sclerosis^22, 23^. An acute increase in serum NfL levels also serves as a highly-predictive marker for poor long-term neurologic outcomes after cardiac arrest^24, 25^. More recently, NfL was investigated in a less strictly neuronal injury-specific context, and attention was drawn to the predictive ability of Nfl with regard to all-cause mortality. Some studies have linked elevated levels of NfL with a higher risk of mortality after stroke and subarachnoid hemorrhage^26–28^. In the elderly, NfL was found to correlate with results on neuropsychological tests, brain atrophy scores but also mortality^29^. Rising and more variable NfL levels were indicative of an accelerated neuronal injury in a population-based cohort study^30^. NfL was also found to predict all-cause mortality in a dementia-free urban population^31^. In general, CI including overt dementia has been linked to an increased risk of all-cause death^32^. In cardiovascular disease NfL levels were found to be elevated in atrial fibrillation and HF^33, 34^. NfL levels also correlate inversely with renal function^35^, and they also show an inverse correlation with BMI and blood volume^36^.

#### Tau

Tau protein is an intracellular microtubule-associated protein abundant in neurons of the CNS and related to axonal transport. It is mostly expressed in cortical neurons but smaller amounts can also be found in other cells, e.g. glial cells. As opposed to NfL, which is highly sensitive for neuronal damage but unspecific to its origins, t-tau is used as a marker of CNS degeneration. It is elevated in many CNS diseases, including stroke, Parkinson’s or multiple sclerosis^37–39^. High CNS t-tau levels in AD patients are indicative of a more rapid cognitive decline and increased mortality^40^. Elevated plasma t-tau levels predicted the risk of incident dementia and stroke in a community-based cohort^39, 41^. T-tau has not been extensively studied in HF populations, but a smaller trial found that it was associated with NT-proBNP in a similar way as found in this cohort^42^. Yet, tau might play a more central role in HF pathology as aggregates were found in healthy and diseased hearts and hyperphosphorylation of tau occurs in HF similar to what is seen in AD^43^.

#### Amyloid beta peptides

The amyloid beta subforms Aβ42 and Aβ40 are the main components of neurotoxic amyloid-β aggregates seen in AD. Aβ peptides are continuously derived from the cleavage of the amyloid precursor protein, an integral membrane protein expressed in many tissues. It is believed that an imbalance between cleavage and degradation results in the accumulation of Aβ peptides in the blood, tissues, and vessels^44^. While Aβ42 is mainly synthesised in the CNS, Aβ40 is also produced peripherally. A decrease in Aβ42 correlates with the formation of AD plaques and the Aβ42/40 ratio improves the diagnostic performance for AD compared to lone amyloid peptide determination. The plasma Aβ42/40 ratio is similarly a robust peripheral biomarker of the cerebral amyloid pathology^45^. In patients with AD, Aβ40 and Aβ42 could be identified within cardiomyocytes and the cardiac interstitium^18^. In the general population circulating Aβ40 has been associated with elevated levels of NT-proBNP and Troponin-T as well as worse cardiac function and higher risk of new-onset HF^46, 47^. In HF, Aβ40 was not associated with cognitive decline but with increased mortality^48^.

This is so far the largest study investigating the relationship of multiple biomarkers of neurodegenerative processes with outcome in a HFrEF cohort. All biomarkers were closely related to HF severity, reflected by a direct correlation with NT-proBNP and NYHA class, and comorbidity burden. Moreover, all biomarkers were highly significant risk factors for worse survival in univariate, but also multivariate models including NT-proBNP. The prognostic performance was best for NfL, which, strikingly, was similar to that of NT-proBNP, the most recognised and validated risk stratificator in HF. Intriguingly, t-tau was also predictive of all-cause mortality in our cohort, performing only slightly worse than NfL or NTproBNP. Importantly, the predictive properties of t-tau are similarly independent of NT-proBNP or kidney function. Elevated NfL and t-tau levels reflect an ongoing, potentially subclinical, neurodegenerative process. Neurodegeneration in HFrEF could be accelerated due to shared risk factors between CI and HF. But also direct mechanisms based on HF, such as impaired cerebral blood flow, proinflammatory disposition or neurohumoral dysregulation have been suggested to play a role^12^. The steady rise in biomarker levels with more advanced disease argues in favour of direct mechanisms. The independent nature of the association of NfL and mortality from NT-proBNP or renal function suggests the contribution of other factors, such as inflammation or microvascular destruction. Accordingly, regression models seeking to explain NfL and tau levels were of very low predictive value, with R^2^ below 0.20. We take this as further evidence that neurodegeneration in CCS is largely independent of classical risk factors, especially age, and is more based on a separate and potentially modifiable mechanism.

Increasing levels of neuromarkers with comorbidity burden could solely reflect the overall load of neuronal injury in the same way that contribute to progression of HFrEF. However, a causal relation cannot be excluded. Mechanistically, the higher levels of neuromarkers in individuals with T2DM may point towards a significant role of hyperglycemia in neuronal injury. In this context, the influence of SGLT2-Inhibitors remains another unresolved question.

A recent trial in HF patients found that elevated NfL levels were related both to white matter hyperintensities as well as hippocampal atrophy^49^. The former is a sign of chronic microvascular damage while the latter is closely related to AD pathology, suggesting that the elevation could be explained by an interplay of cardiovascular changes and amyloid beta-driven pathology. Little is known about the role of t-tau as a marker of neurodegeneration in patients with HFrEF, but our findings suggest an increasing burden of CNS neurodegeneration in more severe cases of HF. This is also evidenced by multiple studies showing degeneration of the cortex, subcortical nuclei and white matter hyperintensities in patients with HF^50, 51^. In AD, the deposition of amyloid into cerebral plaques leads to a lowering in amyloid beta concentrations in the cerebrospinal fluid^52^. Circulating levels of amyloid beta, however, are more prone to influences by peripheral production and elimination by the kidneys, which could contribute to higher levels observed in HFrEF patients^53^. The increase in Aβ40 and Aβ42 seen in our cohort is likely related to a reduced rate of peripheral degradation in patients with HFrEF, including but not limited to reduced renal clearance. This is supported by the regression analysis performed, where unlike tau or NfL, the models for amyloid levels were moderately predictive and included kidney function and volume status as the most powerful explanators. Very little is known about normal plasma ranges of the Aβ42/Aβ40 ratio in HFrEF, but the results here are similar to a recent study in a group of patients at risk for HF^47^. However, unlike either subform in isolation, the ratio was neither associated to NT-proBNP or NYHA, nor with outcome. Taken together, our findings suggest that AD pathology is not a main driver of the CCS.

The biomarkers studied could be used to identify patients prone to accelerated neurodegeneration in CCS. The crucial question is whether early diagnosis of ongoing significant neurodegeneration or CI and therapeutic interventions may slow or improve cognitive decline. Neurohumoral changes accompanying HFrEF are known to drive disease progression and lead to worse outcomes. The vicious cycle of sympathetic overactivation also induces neurotoxicity^54^. Most of the established HF drugs inhibit these pathways. Whether optimizing treatment of HFrEF has a beneficial effect on neuromarker and cognitive decline remains to be investigated.

### Limitations

This study has several limitations. It was conducted at a single tertiary care center, which primarily focuses on a specific population of stable but moderately-severely affected patients with HFrEF. As a result, the generalisability of the findings to a broader population may be limited. Additionally, although patients with a confirmed diagnosis of dementia were excluded, the lack of systematic neuropsychological testing introduces the possibility of contamination from patients across the dementia spectrum. However, based on our data, it appears that primary neurodegeneration through AD pathology is not a prevalent underlying cause within this particular cohort. Nevertheless, further research is needed to explore the relationship between biomarkers of neurodegeneration and neuropsychological assessments in the context of HFrEF.

## Conclusion

In a cohort of patients with moderate – severe HFrEF, markers of neurodegeneration are readily detectable in plasma. Neuromarker levels correlate with HF severity and comorbidity burden. NfL, t-tau, Aβ42 and Aβ40, but not the Aβ42/Aβ40 ratio, are risk factors for worsening HF, i.e. hospitalisations and death, in multivariate models independent from age or NT-proBNP. The prognostic ability of NfL is remarkable and independent from and even comparable to that of NT-proBNP. Together, this data suggest that neurodegeneration, including CNS degeneration, is directly interwoven with the progression of HF and that the CCS is not driven by an acceleration of underlying AD pathology. Novel blood-based neuromarkers, especially NfL, may identify patients with HFrEF warranting a more focused neurological workup. Whether early diagnosis of accelerated neurodegeneration or optimized treatment in HFrEF could be beneficial in maintaining cognitive function represents a new field for investigation

## Data Availability

The data that support the findings of this study are available from the corresponding author upon reasonable request.

## REFERENCES

1. Jack CR, Albert MS, Knopman DS, et al. Introduction to the recommendations from the National Institute on Aging-Alzheimer’s Association workgroups on diagnostic guidelines for Alzheimer’s disease. Alzheimers Dement. 2011;7:257–262.

2. Livingston G, Huntley J, Sommerlad A, et al. Dementia prevention, intervention, and care: 2020 report of the Lancet Commission. The Lancet. 2020;396:413–446.

3. Sonnen JA, Larson EB, Crane PK, et al. Pathological correlates of dementia in a longitudinal, population-based sample of aging. Ann Neurol. 2007;62:406–413.

4. Schneider JA, Arvanitakis Z, Bang W, Bennett DA. Mixed brain pathologies account for most dementia cases in community-dwelling older persons. Neurology. 2007;69:2197–2204.

5. Boyle PA, Yu L, Wilson RS, Schneider JA, Bennett DA. Relation of neuropathology with cognitive decline among older persons without dementia. Front Aging Neurosci. 2013;5.

6. Tiwari S, Atluri V, Kaushik A, Yndart A, Nair M. Alzheimer’s disease: pathogenesis, diagnostics, and therapeutics. Int J Nanomedicine. 2019;Volume 14:5541–5554.

7. Long JM, Holtzman DM. Alzheimer Disease: An Update on Pathobiology and Treatment Strategies. Cell. 2019;179:312–339.

8. Hammond CA, Blades NJ, Chaudhry SI, et al. Long-Term Cognitive Decline After Newly Diagnosed Heart Failure: Longitudinal Analysis in the CHS (Cardiovascular Health Study). Circ Heart Fail. 2018;11:e004476.

9. Hajduk A, Lemon, McManus, et al. Cognitive impairment and self-care in heart failure. Clin Epidemiol. 2013:407.

10. Zuccalà G, Pedone C, Cesari M, et al. The effects of cognitive impairment on mortality among hospitalized patients with heart failure. Am J Med. 2003;115:97–103.

11. Lawson CA, Zaccardi F, Squire I, et al. Risk Factors for Heart Failure: 20-Year Population-Based Trends by Sex, Socioeconomic Status, and Ethnicity. Circ Heart Fail. 2020;13:e006472.

12. Havakuk O, King KS, Grazette L, et al. Heart Failure-Induced Brain Injury. J Am Coll Cardiol. 2017;69:1609–1616.

13. Velliquette RA, O’Connor T, Vassar R. Energy Inhibition Elevates β-Secretase Levels and Activity and Is Potentially Amyloidogenic in APP Transgenic Mice: Possible Early Events in Alzheimer’s Disease Pathogenesis. J Neurosci. 2005;25:10874–10883.

14. Ishizaki T, Erickson A, Kuric E, et al. The Asparaginyl Endopeptidase Legumain after Experimental Stroke. J Cereb Blood Flow Metab. 2010;30:1756–1766.

15. Giannitsi S, Maria B, Bechlioulis A, Naka K. Endothelial dysfunction and heart failure: A review of the existing bibliography with emphasis on flow mediated dilation. JRSM Cardiovasc Dis. 2019;8:204800401984304.

16. Okamoto Y, Yamamoto T, Kalaria RN, et al. Cerebral hypoperfusion accelerates cerebral amyloid angiopathy and promotes cortical microinfarcts. Acta Neuropathol (Berl). 2012;123:381–394.

17. Iadecola C. Neurovascular regulation in the normal brain and in Alzheimer’s disease. Nat Rev Neurosci. 2004;5:347–360.

18. Troncone L, Luciani M, Coggins M, et al. Aβ Amyloid Pathology Affects the Hearts of Patients With Alzheimer’s Disease. J Am Coll Cardiol. 2016;68:2395–2407.

19. Rissin DM, Kan CW, Campbell TG, et al. Single-molecule enzyme-linked immunosorbent assay detects serum proteins at subfemtomolar concentrations. Nat Biotechnol. 2010;28:595–599.

20. Kuhle J, Barro C, Andreasson U, et al. Comparison of three analytical platforms for quantification of the neurofilament light chain in blood samples: ELISA, electrochemiluminescence immunoassay and Simoa. Clin Chem Lab Med CCLM. 2016;54:1655–1661.

21. McDonagh TA, Metra M, Adamo M, et al. 2021 ESC Guidelines for the diagnosis and treatment of acute and chronic heart failure. Eur Heart J. 2021;42:3599–3726.

22. Dominantly Inherited Alzheimer Network, Preische O, Schultz SA, et al. Serum neurofilament dynamics predicts neurodegeneration and clinical progression in presymptomatic Alzheimer’s disease. Nat Med. 2019;25:277–283.

23. Benkert P, Meier S, Schaedelin S, et al. Serum neurofilament light chain for individual prognostication of disease activity in people with multiple sclerosis: a retrospective modelling and validation study. Lancet Neurol. 2022;21:246–257.

24. Moseby-Knappe M, Mattsson N, Nielsen N, et al. Serum Neurofilament Light Chain for Prognosis of Outcome After Cardiac Arrest. JAMA Neurol. 2019;76:64.

25. Wurm R, Arfsten H, Muqaku B, et al. Prediction of Neurological Recovery After Cardiac Arrest Using Neurofilament Light Chain is Improved by a Proteomics-Based Multimarker Panel. Neurocrit Care. 2022;36:434–440.

26. Gendron TF, Badi MK, Heckman MG, et al. Plasma neurofilament light predicts mortality in patients with stroke. Sci Transl Med. 2020;12:eaay1913.

27. Uphaus T, Bittner S, Gröschel S, et al. NfL (Neurofilament Light Chain) Levels as a Predictive Marker for Long-Term Outcome After Ischemic Stroke. Stroke. 2019;50:3077– 3084.

28. Hviid CVB, Gyldenholm T, Lauridsen SV, Hjort N, Hvas A-M, Parkner T. Plasma neurofilament light chain is associated with mortality after spontaneous intracerebral hemorrhage. Clin Chem Lab Med CCLM. 2020;58:261–267.

29. Rübsamen N, Maceski A, Leppert D, et al. Serum neurofilament light and tau as prognostic markers for all-cause mortality in the elderly general population—an analysis from the MEMO study. BMC Med. 2021;19:38.

30. Khalil M, Pirpamer L, Hofer E, et al. Serum neurofilament light levels in normal aging and their association with morphologic brain changes. Nat Commun. 2020;11:812.

31. Beydoun MA, Noren Hooten N, Weiss J, et al. Plasma neurofilament light and its association with all-cause mortality risk among urban middle-aged men and women. BMC Med. 2022;20:218.

32. Dewey ME, Saz P. Dementia, cognitive impairment and mortality in persons aged 65 and over living in the community: a systematic review of the literature. Int J Geriatr Psychiatry. 2001;16:751–761.

33. Sjölin K, Aulin J, Wallentin L, et al. Serum Neurofilament Light Chain in Patients With Atrial Fibrillation. J Am Heart Assoc. 2022;11:e025910.

34. Hoyer-Kimura C, Konhilas J, Sweitzer N, Ryan L, Hay M. Circulating neurodegeneration biomarkers neurofilament light protein and p-tau181 are increased in individuals with heart failure at risk for vascular cognitive impairment. Alzheimers Dement. 2022;18.

35. Akamine S, Marutani N, Kanayama D, et al. Renal function is associated with blood neurofilament light chain level in older adults. Sci Rep. 2020;10:20350.

36. Manouchehrinia A, Piehl F, Hillert J, et al. Confounding effect of blood volume and body mass index on blood neurofilament light chain levels. Ann Clin Transl Neurol. 2020;7:139– 143.

37. Frederiksen J, Kristensen K, Bahl J, Christiansen M. Tau protein: a possible prognostic factor in optic neuritis and multiple sclerosis. Mult Scler J. 2012;18:592–599.

38. Lin W, Shaw J, Cheng F, Chen P. Plasma total tau predicts executive dysfunction in Parkinson’s disease. Acta Neurol Scand. 2022;145:30–37.

39. Pase MP, Himali JJ, Aparicio HJ, et al. Plasma total-tau as a biomarker of stroke risk in the community. Ann Neurol. 2019;86:463–467.

40. Degerman Gunnarsson M, Lannfelt L, Ingelsson M, Basun H, Kilander L. High Tau Levels in Cerebrospinal Fluid Predict Rapid Decline and Increased Dementia Mortality in Alzheimer’s Disease. Dement Geriatr Cogn Disord. 2014;37:196–206.

41. Pase MP, Beiser AS, Himali JJ, et al. Assessment of Plasma Total Tau Level as a Predictive Biomarker for Dementia and Related Endophenotypes. JAMA Neurol. 2019;76:598.

42. Lahiri S, Mastali M, Van Eyk JE, et al. Plasma brain injury markers are associated with volume status but not muscle health in heart failure patients. Front Drug Discov. 2022;2:1042737.

43. Luciani M, Montalbano M, Troncone L, et al. Big tau aggregation disrupts microtubule tyrosination and causes myocardial diastolic dysfunction: from discovery to therapy. Eur Heart J. 2023;44:1560–1570.

44. Mawuenyega KG, Sigurdson W, Ovod V, et al. Decreased Clearance of CNS β-Amyloid in Alzheimer’s Disease. Science. 2010;330:1774–1774.

45. Schindler SE, Bollinger JG, Ovod V, et al. High-precision plasma β-amyloid 42/40 predicts current and future brain amyloidosis. Neurology. 2019;93:e1647–e1659.

46. Stamatelopoulos K, Pol CJ, Ayers C, et al. Amyloid-Beta (1-40) Peptide and Subclinical Cardiovascular Disease. J Am Coll Cardiol. 2018;72:1060–1061.

47. Zhu F, Wolters FJ, Yaqub A, et al. Plasma Amyloid-β in Relation to Cardiac Function and Risk of Heart Failure in General Population. JACC Heart Fail. 2023;11:93–102.

48. Bayes-Genis A, Barallat J, De Antonio M, et al. Bloodstream Amyloid-beta (1-40) Peptide, Cognition, and Outcomes in Heart Failure. Rev Esp Cardiol Engl Ed. 2017;70:924–932.

49. Traub J, Otto M, Sell R, et al. Serum phosphorylated tau protein 181 and neurofilament light chain in cognitively impaired heart failure patients. Alzheimers Res Ther. 2022;14:149.

50. Mueller K, Thiel F, Beutner F, et al. Brain Damage With Heart Failure: Cardiac Biomarker Alterations and Gray Matter Decline. Circ Res. 2020;126:750–764.

51. Stegmann T, Chu ML, Witte VA, et al. Heart failure is independently associated with white matter lesions: insights from the population-based LIFE-Adult Study. ESC Heart Fail. 2021;8:697–704.

52. Andreasen N, Hesse C, Davidsson P, et al. Cerebrospinal Fluid β-Amyloid(1-42) in Alzheimer Disease: Differences Between Early- and Late-Onset Alzheimer Disease and Stability During the Course of Disease. Arch Neurol. 1999;56:673.

53. Arvanitakis Z, Lucas JA, Younkin LH, Younkin SG, Graff-Radford NR. Serum Creatinine Levels Correlate With Plasma Amyloid β Protein: Alzheimer Dis Assoc Disord. 2002;16:187–190.

54. Hartupee J, Mann DL. Neurohormonal activation in heart failure with reduced ejection fraction. Nat Rev Cardiol. 2017;14:30–38.

